# Efficient stochastic epidemic simulation via the Sellke construction

**DOI:** 10.64898/2026.07.16.26358219

**Authors:** Martin C.J. Bootsma, Michiel van Boven

**Author notes:** **Author for correspondence:** Martin Bootsma and Michiel van Boven.

## Abstract

Stochastic epidemic models are a cornerstone of infectious disease epidemiology and are often used to study intervention scenarios. However, large run-to-run variability can make intervention effects difficult to estimate precisely. We revisit the epidemic Sellke construction, which assigns each individual an infection threshold for the cumulative infection hazard such that, conditional on the thresholds, the epidemic trajectory becomes deterministic. This enables coupling of simulations with and without an intervention, yielding low-variance effect estimates even when outcomes such as final size or peak incidence vary widely between runs. We develop an exact, event-driven implementation that maintains infection and recovery events in priority queues. Cumulative infection-hazard updates require *O*(log *N* ) time per event, yielding overall complexity *O*(*E* log *N* ) for *E* events in a population of size *N*. The implementation achieves computational performance comparable to the classical Gillespie algorithm while naturally accommodating non-Markovian infectious periods and complex infectiousness profiles. We illustrate the approach using distance-dependent spread of avian influenza between poultry farms in the Netherlands and a multilayer population with households, schools, and workplaces. In both examples, coupling enables efficient within-run comparisons of intervention scenarios across stochastic realisations.

## 1. Introduction

Stochastic epidemic models are indispensable for studying infectious-disease dynamics in finite populations, and for evaluating interventions such as vaccination, contact reduction, or contact tracing [1]. Unlike deterministic models, stochastic models incorporate random variation in transmission and recovery, which is essential when populations are small or when taking early stochastic extinction into account. At the same time, the variability that makes stochastic models realistic can make them difficult to analyse, in particular when the goal is to quantify the impact of an intervention.

To be precise, a common practical problem is that the outcomes of interest, such as the final size, peak incidence, or epidemic duration, may vary substantially between runs, even when all parameters are fixed [2–4]. When the average effect of an intervention is small relative to the run-to-run variability, large numbers of simulations are required to detect differences reliably. This motivates variance-reduction strategies based on pairing of simulations [5, 6]. Hence, rather than comparing two sets of independent simulations (e.g., without and with intervention), one would like to compare matched realisations that share as much underlying randomness as possible, to isolate the impact of the intervention.

The Sellke construction provides a natural way to achieve such pairing. In this construction, each initially susceptible individual is assigned an exponentially distributed threshold for the cumulative infection hazard (also known as the cumulative force of infection), and infection occurs when that cumulative hazard exceeds the threshold [7, 8]. Conditional on the collection of thresholds and other pre-drawn individual-level random variables such as the duration of the latent and infectious periods, the epidemic realisation then has become deterministic. In fact, the randomness is moved into fixed individual attributes, and the subsequent dynamics is driven by deterministic event times determined by the attributes. The Sellke construction has a long history in epidemic theory, in particular for the calculation of final size distributions [7–9].

With the Sellke construction, one can run paired simulations, for instance without and with an intervention. The resulting coupled outcomes are typically strongly correlated. As a result, the differences between the paired outcomes often have strongly reduced variance compared with an unpaired comparison. In settings where interventions change transmission intensities only moderately (e.g., small changes in vaccination coverage, incremental contact reductions, or targeted measures affecting a subset of contacts), such coupling has the potential to strongly reduce the number of runs needed to achieve a given statistical precision.

Next to coupling, the Sellke framework also separates the infection mechanism (hazard accumulation) from assumptions about infectious-period distributions and infectiousness profiles. Whereas the classical Gillespie simulation algorithm is most direct for Markovian models with exponentially distributed waiting times [10], many applications require non-exponential infectious periods, time-varying infectiousness, or additional individual-level structures [1, 11]. Such features can be handled with ease within the Sellke construction by pre-drawing relevant random variables, without relying on phase-type approximations solely to preserve the Markovian assumptions.

In this paper we investigate the computational performance of Sellke-based simulations relative to those based on the classical Gillespie algorithm [10]. We develop efficient event-driven implementations based on priority queues. The main idea is to maintain candidate infection and recovery events in heaps, and to update cumulative infection hazards in logarithmic time per event, yielding overall complexity on the order of *O*(*E* log *N* ) for *E* events in a population of size *N* . We show that for standard SIR-type models the approach based on the Sellke construction can attain performance comparable to Gillespie simulations, while being much more efficient when assessing the impact of interventions. In addition and as stated above, the Sellke construction allows for more modelling freedom.

We benchmark both approaches across four representative contact structures, namely a complete graph with random mixing, a complete graph with weighted edges, and two sparse graphs, viz. a random regular graph and a small-world great circle network. We then illustrate these ideas in two examples that represent relevant applications of stochastic epidemics: (i) distance-dependent transmission between poultry farms of avian influenza A in the Netherlands [12, 13], and (ii) epidemic transmission on multilayer contact structures representing households, schools, and workplaces.

## 2. Methods

### 2.1 The Sellke construction

Consider a susceptible individual who is exposed to a force of infection *λ*(*t*) at time *t. λ*(*t*) is assumed to be a non-negative, bounded, left-continuous function with at most a finite number of discontinuities. The force of infection *λ*(*t*), or hazard in statistical parlance, can be interpreted as the instantaneous infection risk if the infection has not occurred before time *t*, i.e., in the limit that *δt* ↓ 0, the probability to get infected in the time-interval [*t, t* + *δt*) given that no infection occurred in (−∞, *t*) is given by

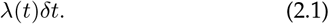

Note that *λ*(*t*) will be continuous on the interval [*t, t* + *δt*) if *δt* is sufficiently small.

The Sellke construction provides an alternative representation of the infection process. Each susceptible individual is assigned an exponentially distributed infection threshold *Q*, with *Q* ∼ Exp(1). Infection occurs when the cumulative force of infection experienced by that individual exceeds this threshold, i.e.,

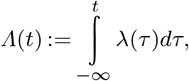

reaches the threshold *Q*. The probability of no infection in the time-interval [*t, t* + *δt*) given that a person was still susceptible at time *t* is given by:

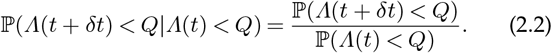

Because *Q ∼ Exp*(1), we have ℙ(*Q* > *a*) = *e*^−*a*^ for all *a* ≥ 0 and equation (2.2) becomes

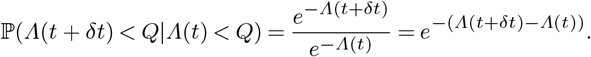

As *Λ*(*t* + *δt*) − *Λ*(*t*) becomes arbitrary small as *δt* ↓ 0, and *λ* is continuous on [*t, t* + *δt*), we can use a Taylor-approximation to write equation (2.2) as

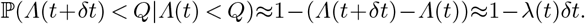

Hence, in the limit that *δt* ↓ 0, the probability that the individual does get infected in the interval [*t, t* + *δt*) given that the individual was susceptible at time *t* is given by

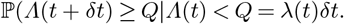

This is identical to equation (2.1). Hence the Sellke construction provides an alternative way to model the infection process [7–9].

The Sellke construction works whenever the infection risk for a susceptible individual can be described by a force of infection. Typically the force of infection experienced by a susceptible individual will consist of the combined contribution of all its infectious contacts. Let *g*_*i*_(*τ* ) be the infectious output of individual *i* where *τ* represents the time since infection and *g*_*i*_(*τ* ) = 0 if *τ* < 0. Let the time-dependent intensity of the contact from individual *i* to individual 1 ≤ *j* ≤ *N* be denoted by *w*_*ji*_(*t*), then we can write the force of infection experienced by individual *j* as

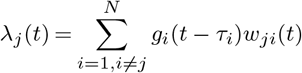

where *τ*_*i*_ is the infection time of individual *i* which is infinite if the individual is not (yet) infected. The time-dependent contact intensity may reflect temporal changes in the network as well as time-dependent infection prevention measures. The Sellke construction, therefore, correctly represents the infection process on any finite, possibly dynamic, graph with weights. Note that one can implement varying susceptibilities of individuals into the threshold, i.e., choose *Q* ∼ *Exp*(*α*) where *α* represents the susceptibility of the individual, but the susceptibilities can also be incorporated into the weights.

## 3. Results

### 3.1 Stochastic coupling

To show why coupling is relevant, we consider the stochastic SIR model with and without vaccination. For large populations without vaccination, the distribution of the final size, conditional on a major outbreak, is normally distributed with mean given by the deterministic final size *Nx*^∗^, where *x*^∗^ is the root of the final size equation 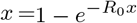 The corresponding variance is given by

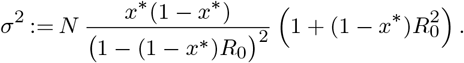

If a proportion *v* of the population is vaccinated with a vaccine that provides perfect immunity, the reproduction number reduces to *R*_*v*_ = (1 − *v*)*R*_0_ and if *R*_*v*_ > 1, conditional on a major outbreak, the final size will again be normally distributed with mean *N* (1 − *v*)*z*^∗^ where *z*^∗^ is the root of

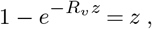

and the variance is given by

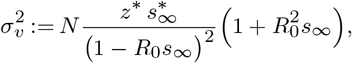

with *s*_*∞*_ = 1 − *v* − *z*^∗^. If we perform *k* independent simulations with and *k* independent unpaired simulations without vaccination the mean difference in the final size will be normally distributed with mean *N* (*x*^∗^ − *z*^∗^) and with variance equal to

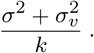

Alternatively, now suppose that we perform *k* paired simulations using the Sellke construction. If *v* is very small, it makes sense to assume that the vaccination has little impact on the size of the outbreak. In the extreme case of perfect correlation between paired runs, the two final sizes can be viewed as small deterministic shifts of the same underlying stochastic realization. In that case, the variance of the paired difference equals

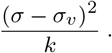

To achieve the same precision for the estimated mean difference in final size, we equate the variances of the two estimators. Let *k*_un_ and k denote the number of simulations in the unpaired and paired designs, respectively. Then,

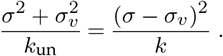

Rearranging this yields

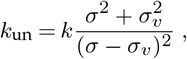

i.e. the number of unpaired simulations must be larger by a factor 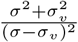 Notice that in realistic settings (*σ* ≠ *σ*_*v*_ ) this factor will be strictly larger than 1. Also note that for small *v* this expression has a leading term

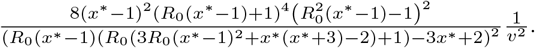

Figure 1 shows this ratio for *R*_0_ = 3, together with numerical results for the stochastic SIR model with *N* = 10, 000, 000. When the effect size becomes of the same magnitude or smaller as the stochastic variations between runs, coupling ensures that far fewer simulations are needed to detect the effect.

**Figure 1.**
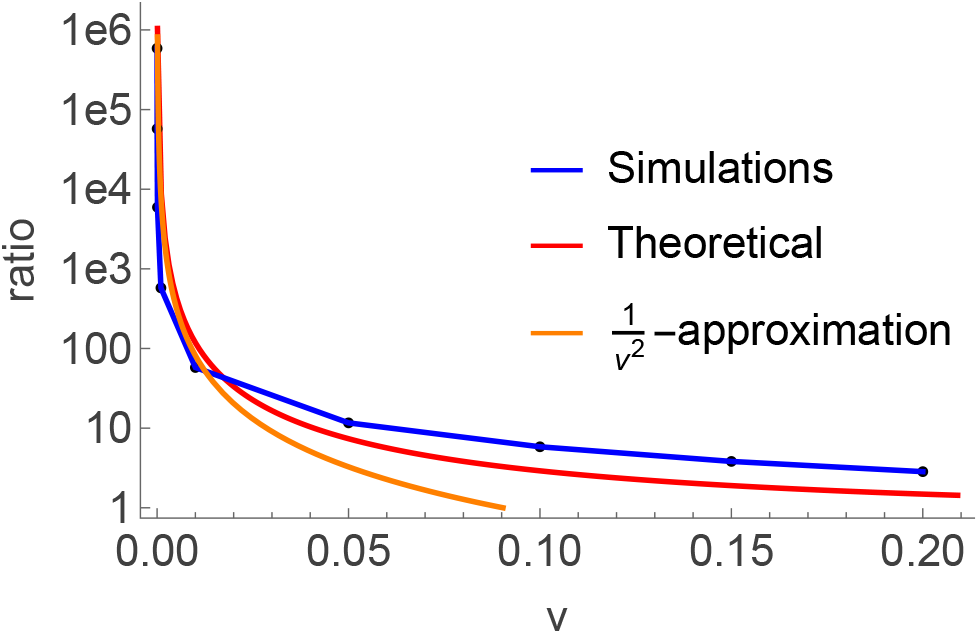
Efficiency gain from coupling. Ratio of unpaired to paired simulations required to achieve equal precision in estimating the intervention effect on final size, as a function of vaccination coverage *v* (*R*_0_ = 3, *N* = 10,000,000). Black dots show numerical estimates from coupled and uncoupled simulations. The orange curve represents the leading-term approximation, and the red curve shows the full analytical expression.

### 3.2 Heap-based implementations

For simplicity, we restrict ourselves to static graphs and develop efficient exact algorithms based on the Sellke construction. At the start of a simulation, we assign (i) an exponentially distributed threshold to each initially susceptible individual and (ii) an infectiousness profile for every individual. For the SIR model with constant infectivity during the infectious period, this reduces to sampling an infectious-period duration for each individual. These quantities are only used if the individual becomes infected. The simulation proceeds as an event-driven algorithm in which infection and recovery events are processed in chronological order.

At each step, the algorithm compares the earliest scheduled recovery time with the earliest time of a susceptible individual crossing the infection threshold. The event with the smaller projected time of occurrence is executed. Because only the earliest recovery time is required, recovery times are stored in a min-heap, which keeps the smallest element at the root of the tree [14]. Both inserting an element into a heap of size *E* and removing the top element take *O*(log *E*) time, while accessing the smallest element takes *O*(1) time.

One advantage of the Sellke construction compared to the Gillespie algorithm [10] is that the Sellke construction requires fewer random numbers. For instance, consider the SIR model on a graph of size *N* with initially one infectious individual. If the final size is *M*, there will have been *M* − 1 infection events and *M* recovery events. For each event, the Gillespie algorithm requires two random numbers, so, in total, 4*M* − 2 random numbers are needed. For the Sellke construction, all (*N* − 1) initially susceptible individuals receive a threshold and all *N* individuals get a duration of the infectious period, provided they get infected. Therefore (2*N* − 1) random numbers are needed. If more than half of the population is infected during the outbreak, i.e. *M* > *N*/2, the Sellke construction requires fewer random numbers. Next we will discuss the implementation on different graphs. In this discussion we ignore time-dependent infection prevention measures. If such measures are introduced, the timing of projected infection events changes and must be handled accordingly.

#### (i) Random mixing

In case of random mixing, all susceptible individuals are exposed to the same infection hazard. Consequently, the first infection event occurs when the cumulative force of infection reaches the smallest remaining threshold. If initially all thresholds are sorted, finding the smallest remaining threshold is easy.

Figure 2 shows the comparative running times for the Gillespie and the Sellke algorithms as function of the population size *N* for the stochastic SIR model, a case tailored for the Gillespie-algorithm. In Figure3, we plot the corresponding ratio of the running times. For the stochastic SIR model, the Gillespie algorithm is a factor 11.2 faster for a population of ten million and the ratio increases with increasing population size as the running time is of order *N* log *N* for the Sellke construction and order *N* for the Gillespie-algorithm. Still, the running time of a single simulation with the Sellke construction takes only minimal time (1.8 seconds in our setting), even in very large populations. Furthermore, the speed of the Sellke construction hardly changes when the infectious period has a non-exponential distribution, while the number of events in Gillespie-algorithm will increase if the infectious period is approximated by a phase-type distribution and the type of the next event will be multinomially distributed [1].

**Figure 2.**
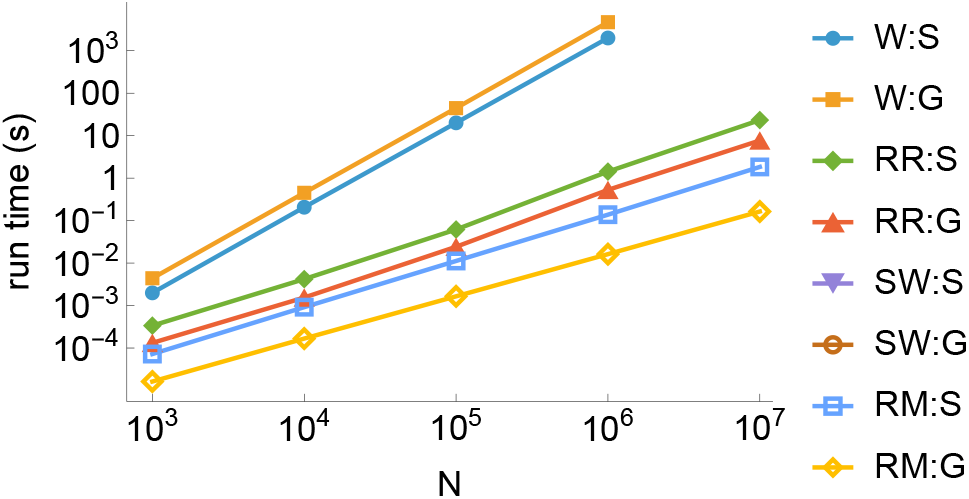
Mean runtime of a single SIR simulation on different contact graphs. We consider a complete graph (random mixing, RM), a small-world (SW) graph (*k* = 12), a random regular (RR) graph (k = 12), and a complete graph with edge weights (W), and compare the computational performance of the Gillespie and Sellke algorithms.

#### (ii) Sparse graphs

In a sparse graph, the average number of neighbours per individual is small compared to the total population size *N* . Infection and recovery events therefore have local rather than global effects, as a change in the infectious status of one individual changes the infection hazards of its susceptible neighbours, but not those of non-neighbouring individuals. Both the Gillespie and Sellke-based algorithms can therefore be implemented using local updating rules (Appendix S2.2).

For the Sellke construction, we store the projected infection times of all susceptible individuals with at least one infectious neighbour in a mutable min-heap. These projected times are the times at which a susceptible individual’s threshold would be reached, conditional on the cumulative exposure already received and on the current set of infectious neighbours. When an individual becomes infected or recovers, only susceptible neighbours whose set of infectious neighbours has changed need to be updated. If a susceptible individual no longer has any infectious neighbours, its projected infection time is removed from the heap, or equivalently set to infinity. Conversely, if a susceptible individual obtains its first infectious neighbour, a projected infection time is inserted into the heap.

For the Gillespie algorithm, one needs to keep track of the set of susceptible-infectious edges, both to compute the total infection hazard and to sample the newly infected individual when the next event is an infection. On sparse graphs with small degree, the number of such edges is of order *N*, and each infection or recovery event requires only local updates. The Gillespie algorithm therefore scales as *O*(*N* ), whereas the Sellke-based algorithm scales as *O*(*N* log *N* ) because projected infection times and recovery times are maintained in heaps. Figure 3 shows the corresponding ratios of running times for a random regular graph and a small-world network. Compared with the random-mixing case, the overhead per infection or recovery event is larger on sparse graphs, because both algorithms must maintain local neighbourhood information. As a result, the relative cost of the heap operations in the Sellke implementation, which gives rise to the logarithmic factor, is less dominant than in the complete-graph case.

**Figure 3.**
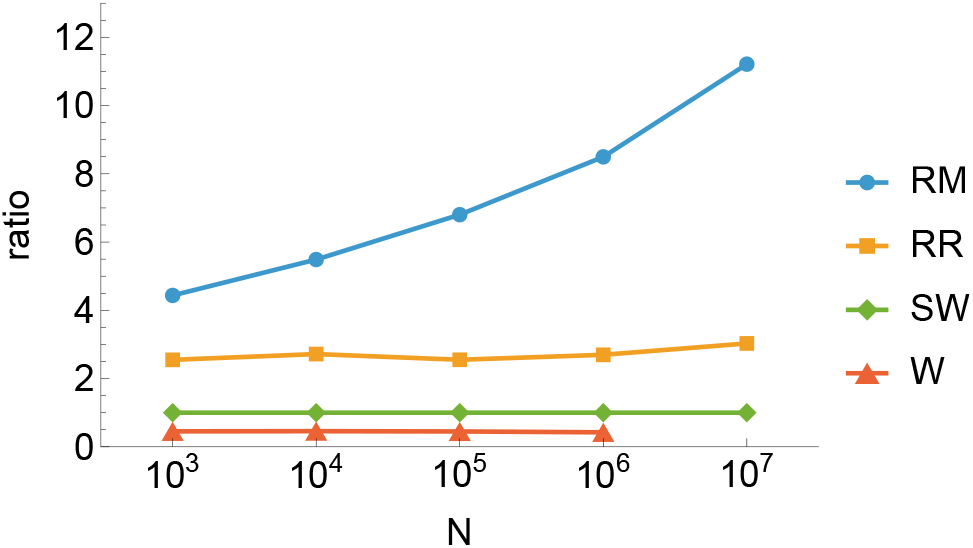
Runtime ratio (Sellke to Gillespie) for an SIR epidemic on different contact graphs of size *N*. We compare a complete graph with random mixing (RM), a small-world (SW) graph (*k* = 12), a random regular (RR) graph (*k* = 12), and a complete graph with edge weights (W).

#### (iii) Complete graphs with weights

For a complete graph with weights *w*_*ij*_ on the edges (1 ≤ *i, j* ≤ *N* and *i* ≠ *j*), the Gillespie algorithm and the Sellke construction are both *O*(*N* ^2^). This can be seen as follows. For the Gillespie algorithm, the total infection rate is 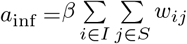 In case of an infection event an edge between a susceptible and an infectious individual is chosen with weight proportional to *w*_*ij*_ . Updating the total infection rate in case of an infection or recovery event is an *O*(*N* ) event. For the Sellke construction, for each infection or recovery event all infection times have to be updated and the minimum of these updated times need to be determined.

Of note, because all projected infection times have to be updated after each event, there is no advantage in storing projected infection events in a heap. Updating all projected infection times and identifying the minimum projected infection time are both *O*(*N* ) operations. Since a simulation contains *O*(*N* ) infection and recovery events, the overall running time is *O*(*N* ^2^). Figure 3 shows the ratio of running times for the case in which all individuals are assigned independent uniform random positions on the unit square and edge weights depend on the distance between the two endpoints.

### 3.3 General infectiousness profiles

Up to this point, we have assumed that the infectiousness of an individual is constant during its infectious period. The Sellke construction can also be applied when individual *i* has a general infectiousness profile *g*_*i*_(*τ* ), with *τ* denoting the time since infection. If there is no finite time *T* such that *g*_*i*_(*τ* ) = 0 for all *τ* > *T*, then recovery is not explicitly modelled and no recovery heap is needed. The disadvantage is that more computational effort is required to determine when a susceptible individual reaches its threshold.

At time *t*, for each susceptible individual *j*, one has to find the calendar time *t*_*j*_ at which the current set of infected neighbours of *j* will have produced enough infectious pressure for the threshold *Q*_*j*_ to be exceeded, i.e.,

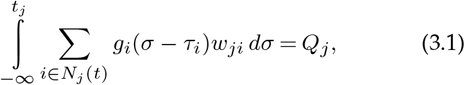

where *N*_*j*_ (*t*) is the set of neighbours of individual *j* that were infected before time *t, τ*_*i*_ is the infection time of individual *i*, and *Q*_*j*_ is the Sellke threshold of individual *j*. Here, *g*_*i*_(*τ* ) = 0 for *τ* < 0. The current set of infected neighbours *N*_*j*_ (*t*) is insufficient to cause infection if

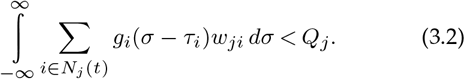

As long as the set of infected neighbours of individual *j* does not change, the projected infection time *t*_*j*_ remains unchanged. Once individual *j* has a finite projected infection time, this time can only move earlier when additional neighbours become infected.

### 3.4 Generation-based algorithm

If only the final outbreak size is of interest, the Sellke construction allows for an efficient generation-based algorithm [15–18]. An attractive feature of the Sellke construction is that a simulation can switch from the time-based event-driven algorithm to a generation-based algorithm once infection times are no longer required. This can substantially reduce running time, because exact infection and recovery times no longer need to be computed. We have used this strategy in the avian influenza example below.

### 3.5 Example: Complete weighted graphs

Our first example considers the transmission of avian influenza between poultry farms in the Netherlands [12, 19–21]. This provides a challenging example for the algorithms developed above, because the contact graph is complete and weighted. Specifically, each farm can transmit infection to every other farm, with a pairwise transmission hazard that depends on the distance between the two farms. Thus, unlike in sparse graphs, local updating is not feasible after an infection or recovery event.

In the example, poultry farms are classified as susceptible, infected, or recovered and immune. The transmission hazard from infected farm *i* to susceptible farm *j* is proportional to a distance-dependent kernel evaluated at the Euclidean distance between the two farms. As in earlier analyses, we use a three-parameter logistic function for this kernel and calibrate the overall transmission scale such that the basic reproduction number has the desired value in an infinitely large spatially homogeneous population [12]. Compared with earlier analyses, we use updated anonymised farm locations corresponding to the 2026 census [22]. The data consist of 2, 028 commercial poultry farms.

For each farm in turn, we initiate an epidemic with that farm as the index case. For each index farm, we generate 100 paired realisations, using the same Sellke thresholds and infectious period realisations for the simulation without intervention and the corresponding simulation with intervention. In the intervention scenario, the three geographically nearest neighbouring farms of the index farm are removed before the start of the simulation. This mimics a highly localised pre-emptive culling strategy around the index premises. The setup is related to earlier work on risk-based culling, in which farms were prioritised according to their expected contribution to onward transmission [19].

The pairing of simulations is important. In earlier evaluations of culling strategies, many independent simulations were required, and the outcomes were analysed with a linear mixed model to separate variation due to culling strategy, farm map, and stochastic epidemic realisation [19]. In the present setting, the Sellke construction provides a natural coupling of simulations with and without intervention. Because the paired simulations share the same underlying Sellke thresholds and infectious-period realisations, stochastic variation between baseline and intervention runs is strongly reduced.

The effect of removing the three nearest neighbours of the index farm is highly spatially heterogeneous (Figure 4). For most index farms, the mean reduction in final outbreak size is close to zero. This occurs either because introductions often die out stochastically even without intervention, or because removing three neighbouring farms is too small a perturbation to alter the subsequent epidemic trajectory. Larger reductions are concentrated in poultry-dense areas, where local transmission between nearby farms can sustain epidemic spread. This is consistent with earlier analyses showing that the risk of epidemic spread is concentrated in high-density poultry areas [12].

**Figure 4.**
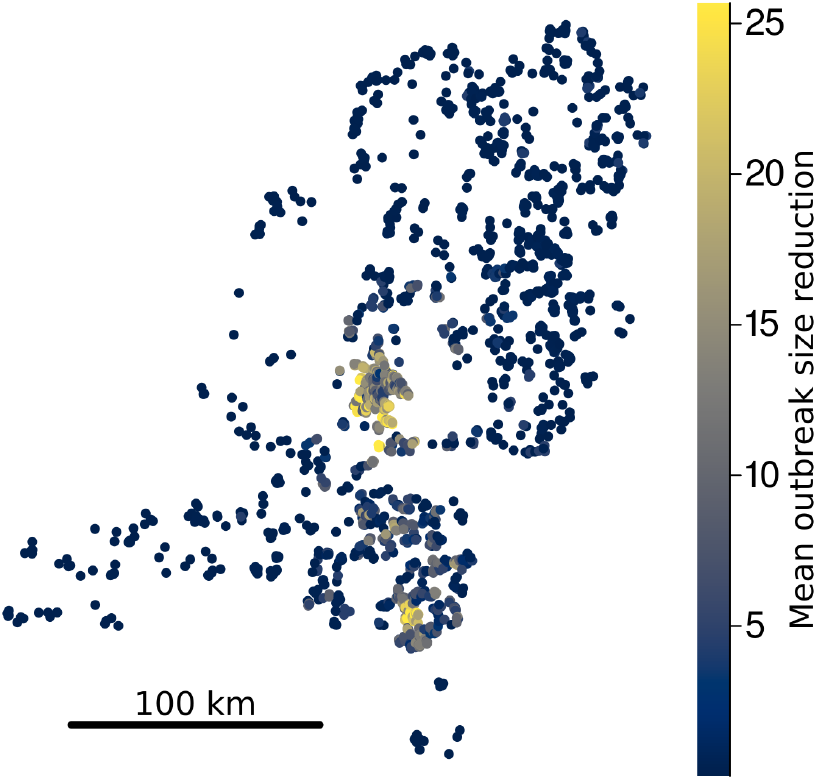
Spatial variation in the effect of local pre-emptive culling in the avian influenza example. Shown are the locations of all 2, 028 commercial poultry farms in the Netherlands. Each farm is used 100 times as the index case of a simulated epidemic, yielding 202, 800 paired intervention comparisons. Colours indicate the mean reduction in final outbreak size when the three geographically nearest neighbouring farms of the index farm are removed before the start of the simulation, compared with the paired simulation without intervention. For privacy reasons, random jitter has been applied to the actual farm locations.

The paired final sizes show that the intervention has a strongly variable effect across stochastic realisations (Figure 5). Many points lie close to the diagonal, indicating that removing the three nearest neighbours often has little effect on the final size. However, there is also a substantial set of simulations far below the diagonal, including simulations in which the outbreak remains very small with intervention while the paired outbreak without intervention becomes large. These cases illustrate why paired simulations are useful as final outbreak sizes vary strongly even for the same index farm, and the effect of a local intervention will be difficult to tease out unless both interventions simulations are paired to simulations without the intervention.

**Figure 5.**
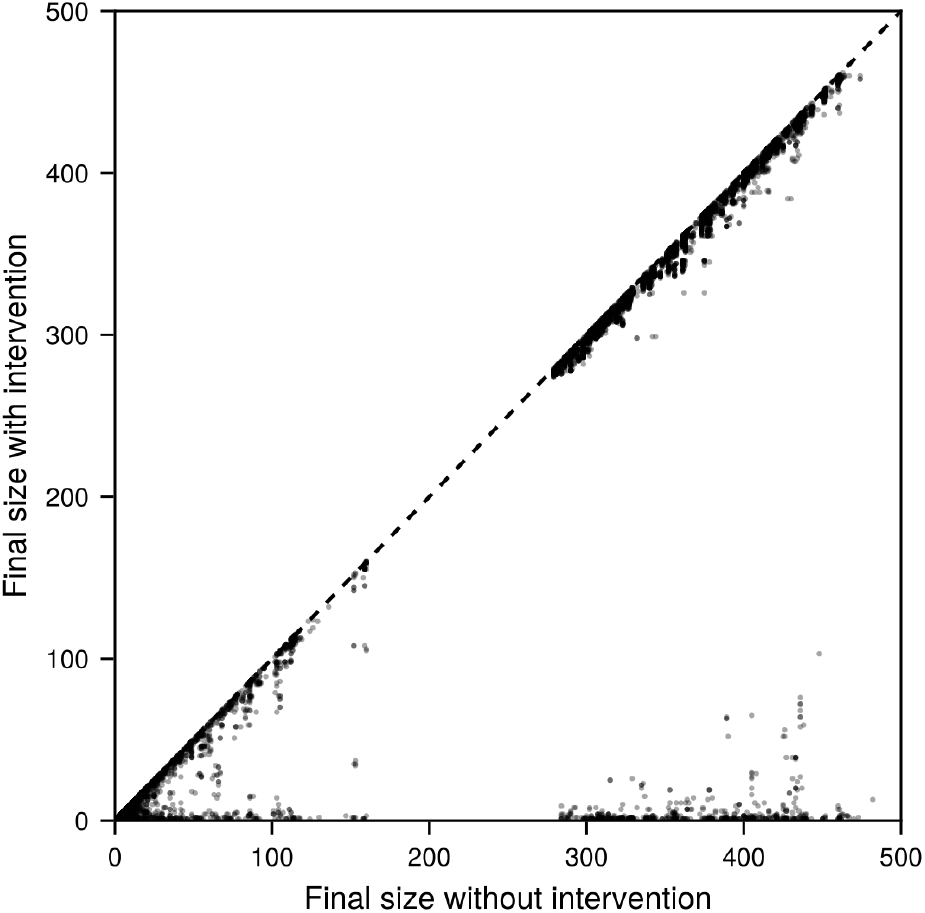
Paired final outbreak sizes in the avian-influenza example. Each point represents one paired simulation, generated using the same Sellke thresholds and infectiousness profiles in the baseline and intervention scenarios. The horizontal axis shows the final outbreak size without intervention, and the vertical axis shows the final outbreak size under the indicated intervention. The dashed line indicates equal final outbreak sizes. Points below the dashed line correspond to simulations in which the intervention reduced the final outbreak size.

### 3.6 Example: Community-structured graphs

Our second example considers a sparse graph based on a community structured into households, schools, and workplaces. Such community-structured contact networks are widely used in individual-based and agent-based transmission models of respiratory viruses, because they represent the main settings in which repeated close contacts occur and allow interventions such as case isolation, household quarantine, school closure, and workplace closure [23–28].

The population consists of 2, 000 households, each with two adults and two children, giving a total population size of 8, 000. Household locations are positioned uniformly on the unit square. Schools are placed on a regular grid, and each child attends the nearest school. Adults are assigned uniformly at random to one of 80 workplaces. Workplaces are not spatially embedded, and their sizes therefore vary because of the random assignment of adults to workplaces. Transmission can occur within households, schools, and workplaces. Household transmission is density dependent, with *β*_HH_ = 0.6, whereas transmission in schools and workplaces is frequency dependent, with *β*_S_ = *β*_WP_ = 0.9 [29]. Notice that the example can readily be adjusted to incorporate actual demographic, spatial, contact, or institutional data.

We compare a baseline scenario without interventions with symptom-triggered interventions. In all intervention scenarios, detected symptomatic individuals no longer contribute to transmission at school or at work, and their infectiousness within the household is reduced by 40%. In addition, we consider household quarantine, school closure, workplace closure, and combinations of these measures. Household quarantine removes all household members from school and workplace transmission for 3.62 days after the most recent detected household case, while school and workplace closures are triggered by a detected case in the corresponding setting and last for a fixed duration of 3.62 days.

The simulations show substantial variation in final outbreak size across stochastic realisations, and in the impact of interventions (Figure 6). Many realisations remain small even without intervention, leaving little room for reduction, whereas for larger baseline outbreaks the reactive interventions can markedly reduce final size, as indicated by points well below the diagonal. The largest reductions occur when the baseline epidemic takes off but the corresponding intervention scenario remains limited. Thus, the paired comparisons show both the strong intrinsic variability of epidemic outcomes and the ability of reactive interventions to prevent some otherwise large outbreaks. Such direct comparisons would have been difficult without coupling the intervention and baseline scenarios.

**Figure 6.**
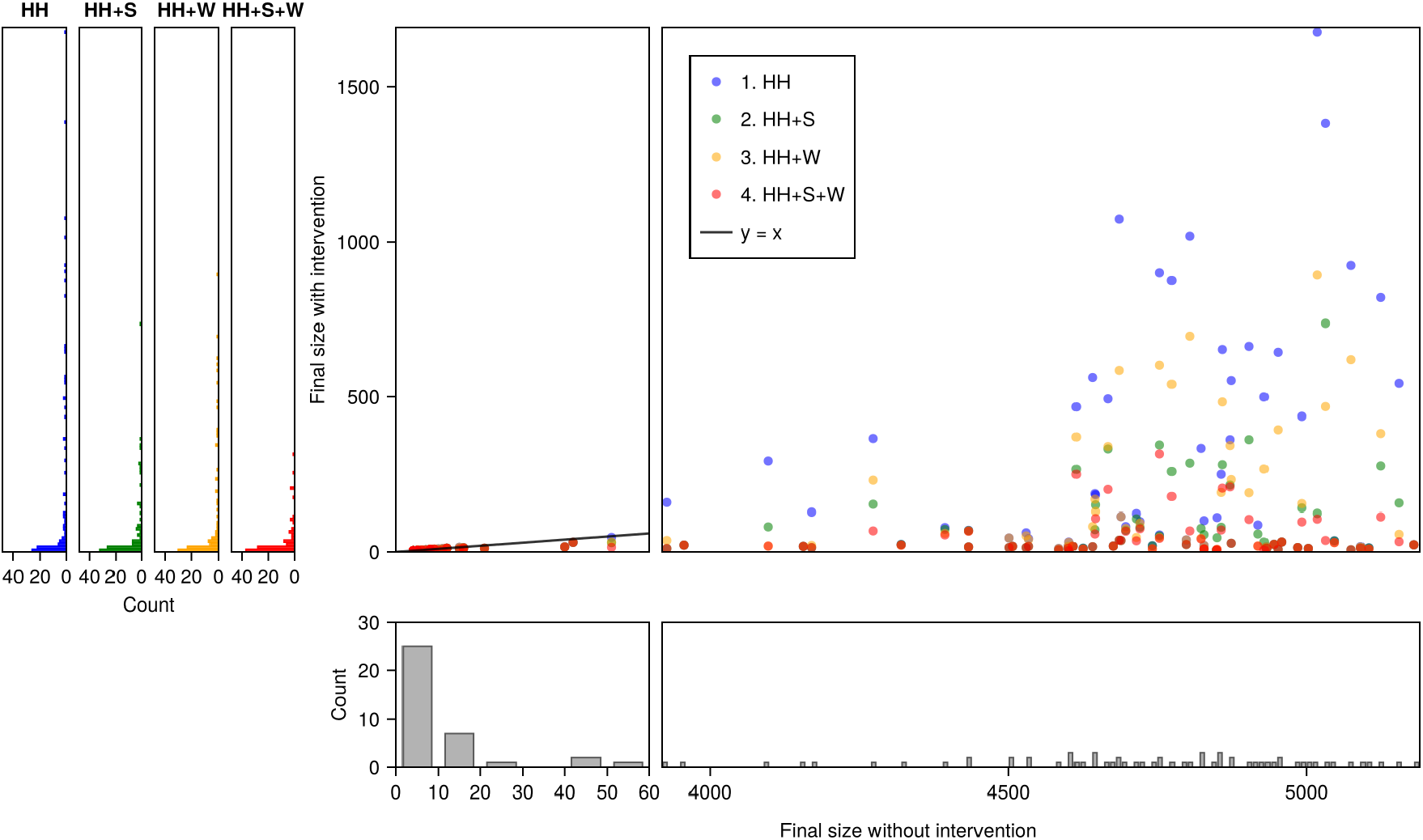
Paired final outbreak sizes under interventions in a community-structured graph. Each point represents one paired simulation, generated with the same Sellke thresholds, infectiousness profiles, and symptom-onset times in the baseline and intervention scenarios. The intervention scenarios are HH, comprising isolation of detected symptomatic individuals and quarantine of their household members; HH+S, adding temporary school closure following detection of a case; HH+W, adding temporary workplace closure following detection of a case; and HH+S+W, adding temporary closure of both schools and workplaces. Household quarantine and school or workplace closure last 3.62 days, corresponding to the time by which 80% of total infectiousness has occurred. The horizontal axis shows the final outbreak size without intervention, and the vertical axis shows the final outbreak size under the indicated intervention. The dashed line indicates equal final outbreak sizes. Points below the dashed line correspond to simulations in which the intervention reduced the final outbreak size.

## 4. Discussion

We developed an efficient event-driven simulation approach for stochastic epidemic models based on the Sellke construction and argued that it has important advantages over the well-known Gillespie algorithm [10]. Specifically, by using heaps to manage infection and recovery events, our algorithm runs in *O*(*E* log *N* ) time for a population of size *N* with *E* events, improving significantly on naive implementations based on linear scans or search-tree–based event lists. In settings tailored to the assumptions of the Gillespie algorithm, the heap-based Sellke simulations are somewhat slower than corresponding simulations based on the Gillespie algorithm (Figures 2-3). However, the ability of the Sellke framework to couple epidemic simulations with and without an intervention produces correlated outcomes and thereby reduces the variability in estimated intervention effects. This allows even subtle differences to be detected with far fewer simulations than would be required with independent simulations. We illustrated these benefits in two case studies: a spatially structured epidemic in a fully connected network model, and a community with layered contacts (households, schools, workplaces). Furthermore, an added benefit of the Sellke-based simulations is that it naturally accommodates model generalisations, including non-exponential latent and infectious period distributions, and complex infectiousness profiles. Recent real-world examples that have relied on the Sellke construction include likelihood-based estimation of individual-level susceptibilities [30] and infectious periods [31], and the modelling of the asymptomatic phase in a multi-scale HIV model [32]. Our study has shown how the computational efficiency of such analyses can be improved.

An additional computational advantage arises when the primary quantity of interest is the final outbreak size rather than the full temporal trajectory. In that case, the Sellke construction permits a generation-based algorithm in which only cumulative infection pressures are propagated forward. Individuals whose residual thresholds are exceeded by the remaining infectious pressure of the current generation form the next generation, and the procedure is repeated until no further infections occur. Because this avoids explicit scheduling and comparison of infection and recovery times, heaps are no longer needed. This connects the computational use of the Sellke construction to classical final-size arguments based on infection generations and to graph-based calculations of outbreak sizes on networks [8, 9, 15, 16, 33].

Whilst we have provided efficient heap-based algorithms to simulate stochastic epidemics, we do not claim that our methods are computationally optimal. Instead, we focused on presenting the main conceptual arguments. Within our setting, several extensions and refinements may further improve computational performance. First, one may exploit structural bounds to infection inherent to the Sellke representation. Specifically, an individual whose resistance threshold exceeds the total infectious pressure that could possibly be exerted by all its infected neighbours can never become infected. Identifying and pruning such individuals from the network at the outset could reduce the effective population size, particularly in settings with strongly heterogeneous degree distributions. Second, the size of the infection heaps can be reduced by avoiding insertion of infection times that cannot occur within the infectious period of the source case. Hence, one may restrict heap insertions to neighbours whose resistance thresholds can be exceeded before recovery of the focal infected individual. In settings with short infectious periods or limited transmission potential, this may substantially limit the number of heap operations. Third, alternative heap implementations may yield additional gains. While binary heaps offer a natural baseline, d-heaps (e.g., 4-heaps), or interval-based heaps (e.g., bucketing per day or week) could potentially further reduce the number of computations and model complexity. The optimal choice depends on the relative frequency of insertions, deletions, and related updates, and may vary with network size and structure. This is a natural direction for further analyses.

An important use of mathematical models is the evaluation of counterfactual scenarios that consider post-hoc evaluation of alternative interventions. In deterministic epidemic models, such analyses are straightforward as alternative interventions can be assessed by modifying model parameters or structure. This yields outcomes that are directly comparable to a baseline scenario. In stochastic epidemic models, however, independent simulations under different interventions are subject to Monte Carlo variability, potentially obscuring the effects of an intervention. Coupling epidemic realisations through shared underlying randomness provides the stochastic analogue of deterministic counterfactual analysis, enabling exact and low-variance comparisons between alternative interventions. The importance of such pairing in stochastic epidemic models has been recognised previously. For instance, using examples from historical epidemics of citrus canker in Florida and common cold on the island of Tristan Da Cunha, matched epidemic histories were constructed to provide post-hoc evaluations of alternative intervention strategies [5]. In a methodologically related paper, a graphical *single-world* was constructed to generate matched counterfactual realisations in a simulation study, thereby eliminating stochastic variability between paired baseline and intervention simulations [6]. The analyses in these papers underline the value of coupling for counterfactual analyses, but focus primarily on intervention evaluation rather than computational efficiency. Specifically, pairing is achieved through explicit constructions of shared potential transmission links, whereas the Sellke framework yields coupling directly through a threshold representation. As we have argued, the Sellke construction, when properly implemented, has favourable scaling properties.

Taken together, our results demonstrate that the Sellke construction provides not only a transparent probabilistic representation of stochastic epidemics, but also forms a computationally efficient basis for large-scale simulation and exact counterfactual analysis. Specifically, the algorithms scale favourably with population size and number of events, making it feasible to simulate complex epidemic models in populations of realistic size (up to millions). This is particularly relevant in settings where the evaluation of multiple sets of interventions is required in near real-time.

## Data Availability

No data are used in this study. All code will be deposited on github.

## Data Accessibility

Code is available at https://github.com/martinbootsma/heapcode.

## Authors’ Contributions

M.C.J.B. and M.v.B. jointly conceived the study, developed the methodology, and wrote the manuscript. M.C.J.B. implemented the simulation code.

## Competing Interests

The authors declare no competing interests.

## Funding

This work was supported by the Netherlands Organisation for Health Research and Development (ZonMw), project *Risk assessment for zoonotic transmission of avian influenza in the Netherlands* (grant number 10150022310017).

## Acknowledgements

Elise van den Broek is gratefully acknowledged for valuable input during the conception of this manuscript. Dr. Thomas Hagenaars and Dr. Gert-Jan Boender of Wageningen Univeristy and Research are gratefully acknowledged for providing the anonymised farm location data.

## Supplementary material

This supplementary material provides algorithmic details, implementation remarks, and pseudocode for the Sellke-based algorithms described in the main text. Our github repository also contains code to runs various instances of the Gillespie algorithm.

## S1. Remarks on heaps

The heap implementations described in this paper are intended primarily to illustrate the algorithms rather than to provide a fully optimal implementation. Several alternative priority-queue data structures could potentially improve computational performance. For example, replacing binary heaps by *d*-ary heaps can reduce the height of the tree and thereby decrease the number of levels traversed during insertions and deletions. Values of *d* around four sometimes are considered a reasonable compromise between the reduced tree depth and the larger number of comparisons required at each level [1]. However, for our specific problems it is likely that the optimal choice will depend on the specific problem at hand, and possibly also on the available hardware.

Another possible optimisation is to partition events across bucketed heaps, with each heap containing event times falling within a specified time interval. This keeps the individual heaps smaller and may reduce the computational cost of insertion and deletion operations. Such an approach is particularly natural for the recovery heap, because recovery times are fixed once inserted. It may also be useful for infection heaps implemented using lazy deletion. For mutable infection heaps, projected infection times may change repeatedly as the infection status of neighbouring individuals changes. In a bucketed implementation, such updates may require elements to be moved between buckets. Whether the reduced heap sizes compensate for this additional bookkeeping is therefore likely to depend on the population size, event density, contact structure, and frequency of updating.

## S2. Pseudocode

### S2.1 Sellke construction for the stochastic SIR model

The homogeneously mixing SIR model provides the simplest implementation of the Sellke construction. Because all susceptible individuals experience the same cumulative infection pressure, their experienced cumulative infection pressures need not be tracked separately. Hence, it is sufficient to order their Sellke thresholds and determine when the next threshold is crossed. Infection events can therefore be calculated directly from the current number of infectious individuals, while recovery times need to be maintained in a priority queue. The resulting algorithm generates the exact stochastic trajectory and avoids repeatedly selecting transmission pairs. Its main computational requirements are generating the ordered thresholds and maintaining the recovery heap. The thresholds may either be sorted explicitly or generated efficiently from their exponentially distributed spacings using Rényi’s representation. Below is pseudocode of the model.

**Require**: Population size *N*, rates *β, γ*, initial counts (*S*_0_, *I*_0_, *R*_0_)

**Ensure** Event-driven trajectory (*t, S, I, R*)

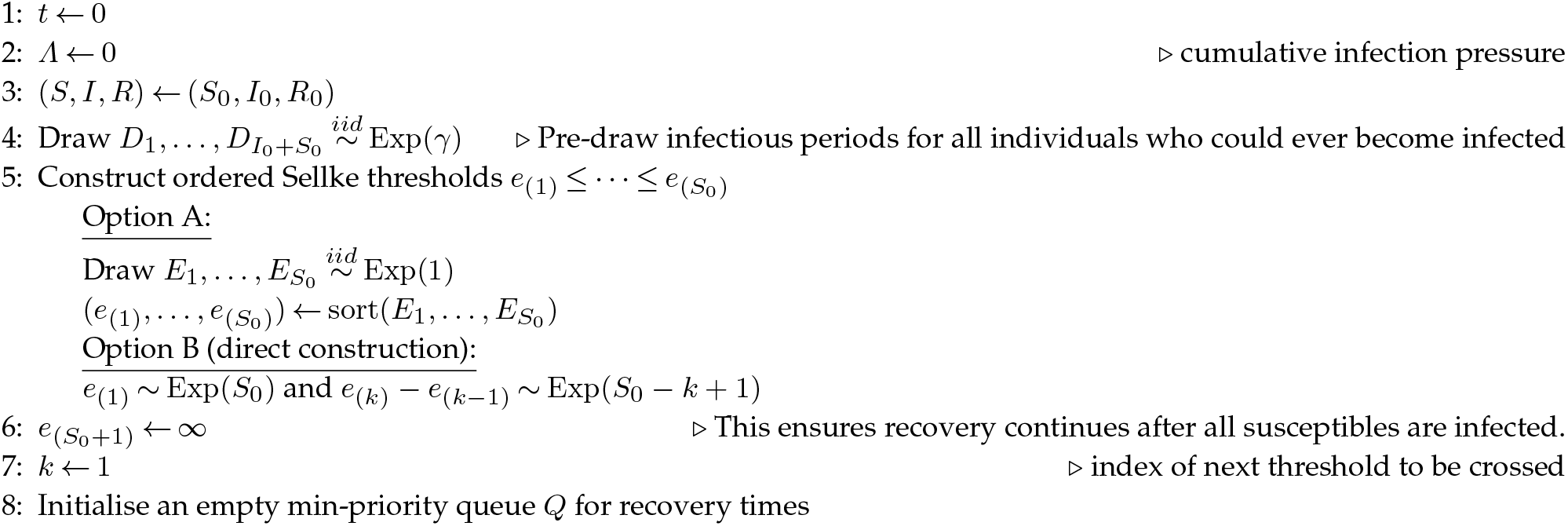
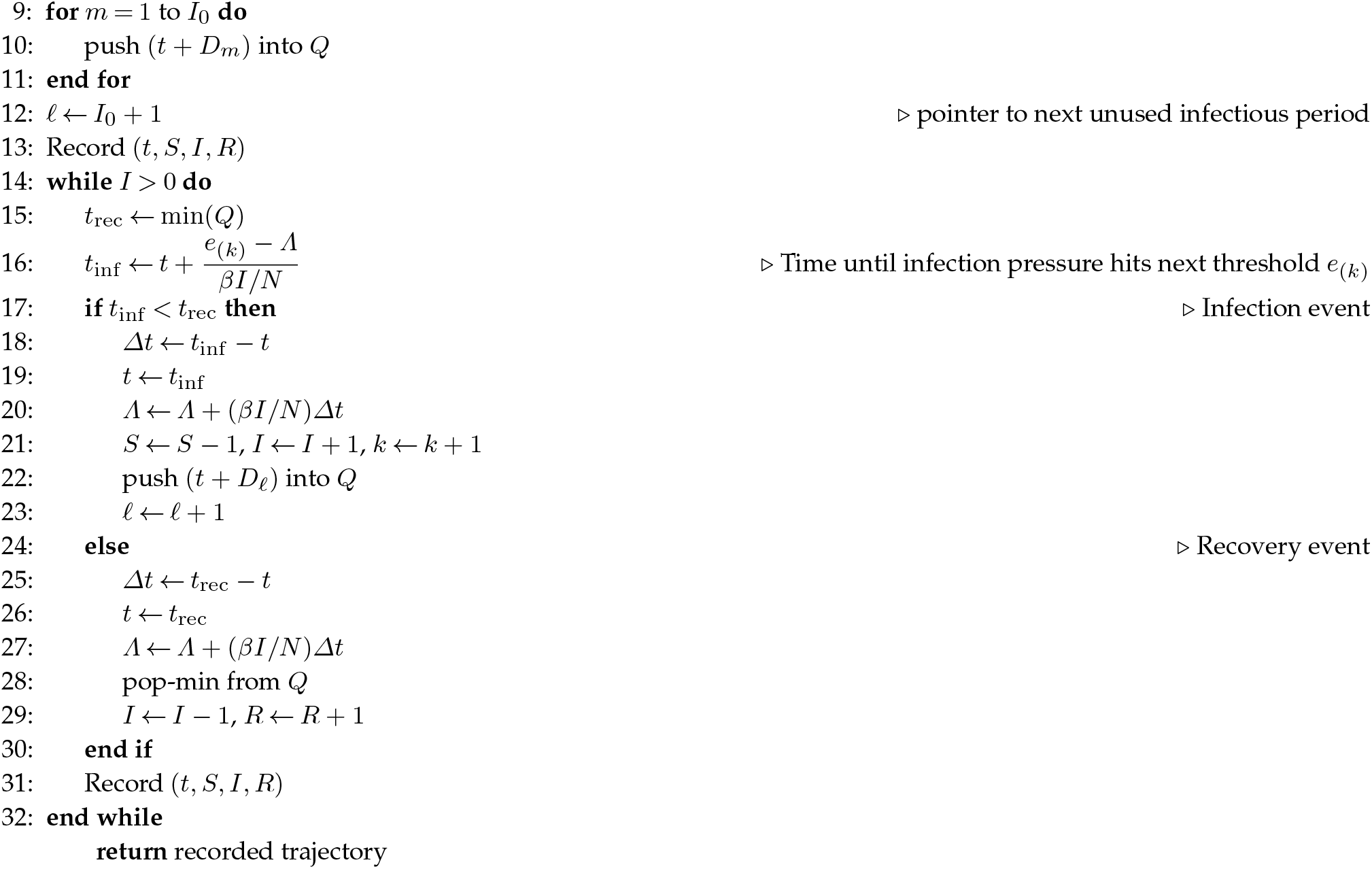

### S2.2 Sellke construction for the stochastic SIR model on a sparse graph

On a sparse graph the cumulative infection pressure is specific for each individual, but changes only for susceptible neighbours of a node that becomes infected or recovers. The algorithm therefore stores, for each susceptible individual, its remaining Sellke resistance and current number of infectious neighbours. These quantities determine a projected infection time, which is maintained in a mutable priority queue, while recovery times are stored in a standard min-heap. The main advantage is that each event requires only local updating, making the algorithm particularly suitable for graphs with small degree relative to the population size. The main implementation complication is that tentative infection times may move both forwards and backwards, and nodes may enter or leave the infection heap. Efficient heap updates are therefore required.

**Require:** Static graph adjacency lists {*N* (*v*)}_*v*∈*V*_, rates *β, γ*, initial infected set *ℐ*_0_, initial recovered set *ℛ*_0_

**Ensure:** Infection times *τ*_*I*_ [*v*] and recovery times *τ*_*R*_[*v*] for all nodes *v*; set *τ*_*I*_ [*v*] = +∞ if *v* is never infected for *t* ≥ 0, and set *τ*_*R*_[*v*] = −∞ for *v* ∈ *R*_0_.

**State:** state[*v*] *∈ {S, I, R}*

**Sellke resistance:** for initially susceptible *v*, draw *E*[*v*] *∼* Exp(1)

**Susceptible bookkeeping:** *c*[*v*] = number of infected neighbors of susceptible *v, r*[*v*] = remaining resistance, *ℓ*[*v*] = last time *r*[*v*] was updated

**Heaps:** *H*_*I*_ = mutable min-heap keyed by tentative infection times *t*_*I*_ [*v*] (stores susceptible nodes; supports DECREASEKEY/INCREASEKEY/DELETE via handles), *HR* = standard min-heap keyed by recovery times *τR*[*v*] (infected nodes)

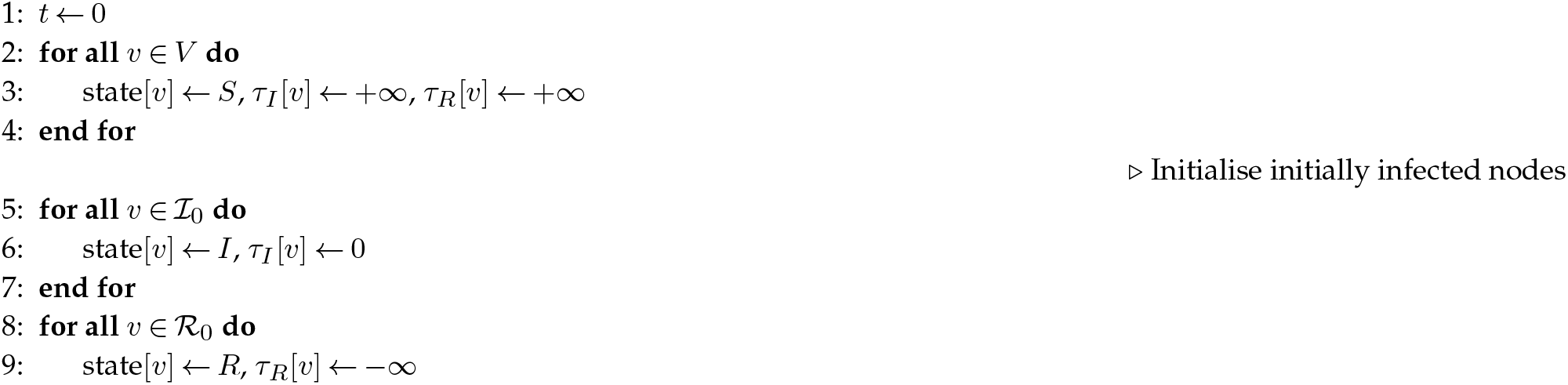
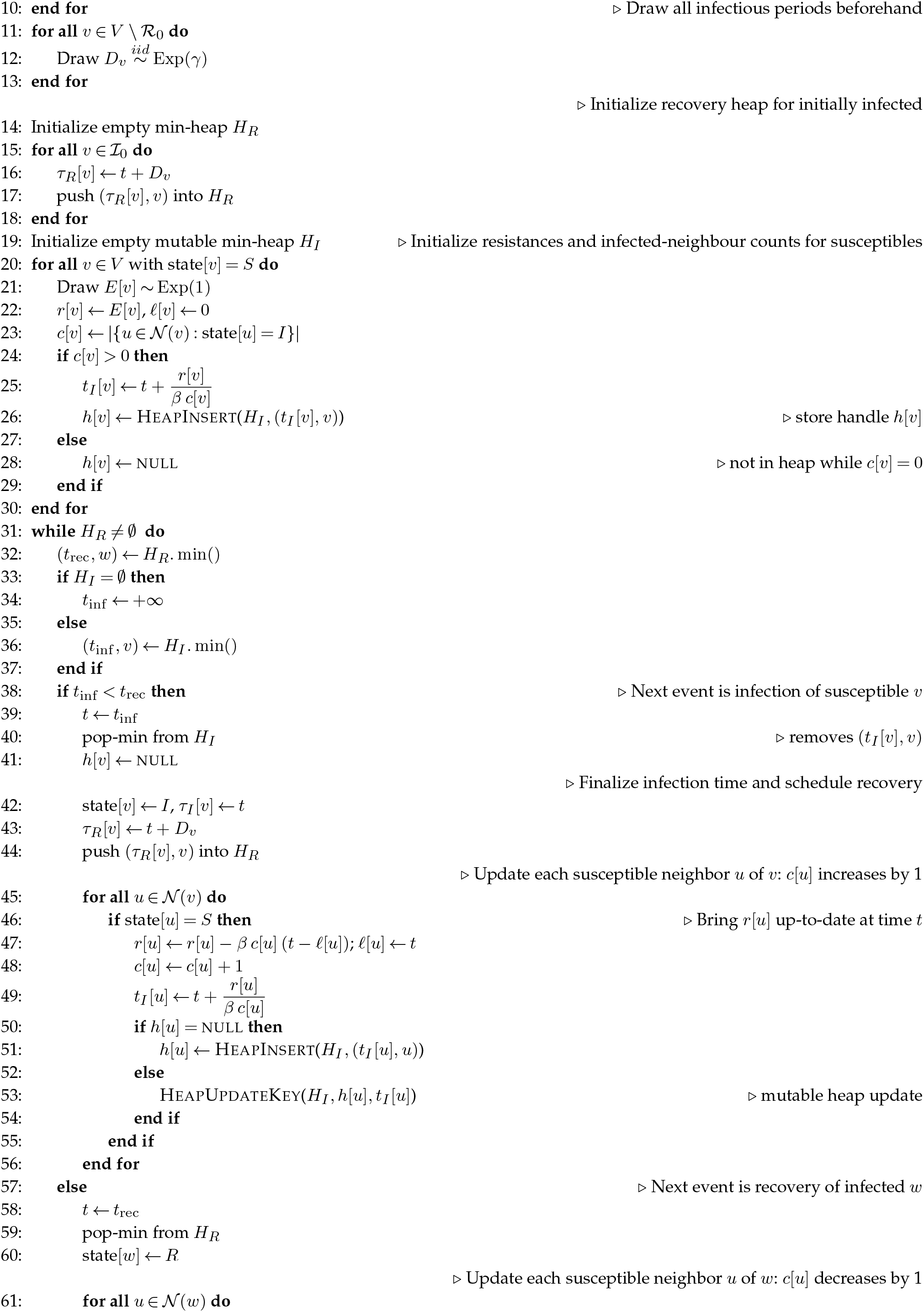
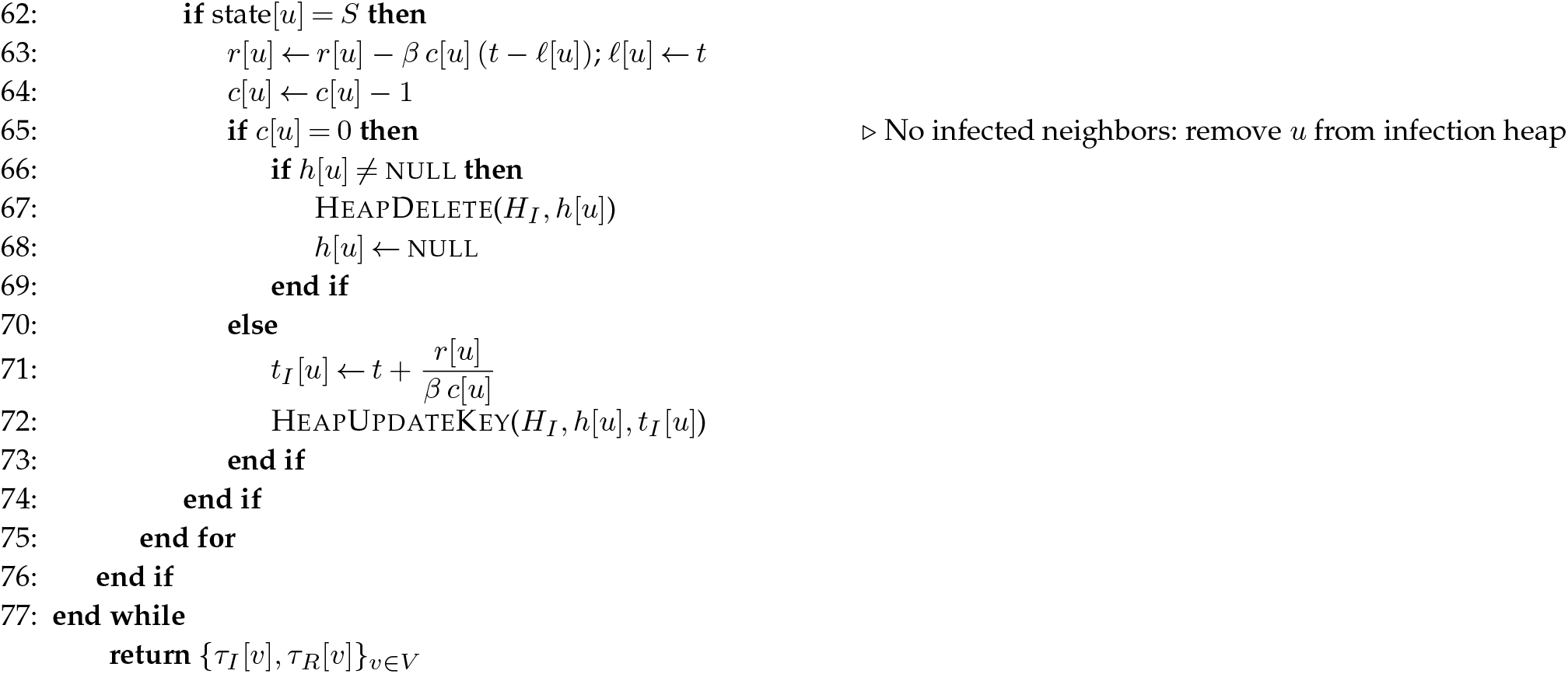

### S2.3 Sellke construction for the stochastic SIR model on a complete weighted graph

For a dense weighted graph, each susceptible individual has its own infection pressure (as in the sparse graph), determined by the weighted contributions of all currently infectious nodes. This formulation accommodates arbitrary pairwise heterogeneity while retaining the exact threshold-crossing interpretation of the Sellke construction. However, an infection or recovery may change the pressure on almost every susceptible node. Maintaining all tentative infection times in a mutable heap would then require updating a large fraction of the heap after each event and offers little computational advantage. The algorithm below instead scans the susceptible nodes to identify the next threshold crossing and updates their infection pressures directly. This entails *O*(*N* )) operations per event and therefore *O*(*N* ^2^) operations over a complete epidemic in the worst case. The algorithm is, however, straightforward, and it avoids the bookkeeping associated with a mutable infection heap.

**Require:** Nodes *V* = *{*1, …, *N}*, weights *w*_*ij*_ ≥ 0 with *w*_*ii*_ = 0, rates *β, γ*, initial infected set ℐ_0_, initial immune/recovered set ℛ_0_

**Ensure:** Infection times *τ*_*I*_ [*v*] and recovery times *τ*_*R*_[*v*] for all *v*; *τ*_*I*_ [*v*] = +*∞* if never infected for *t* ≥ 0, and *τ*_*R*_[*v*] = −∞ for *v* ∈ ℛ_0_

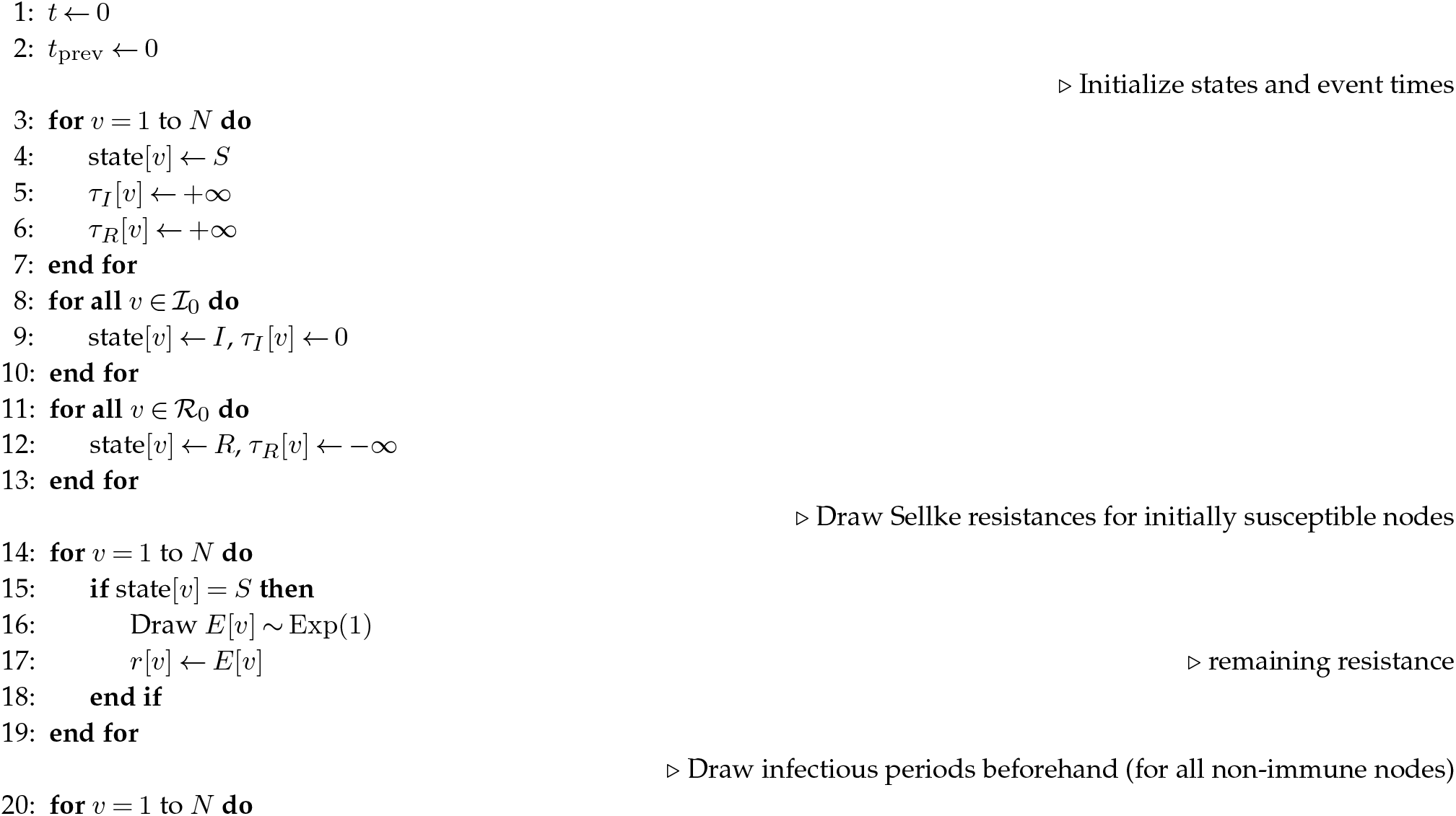
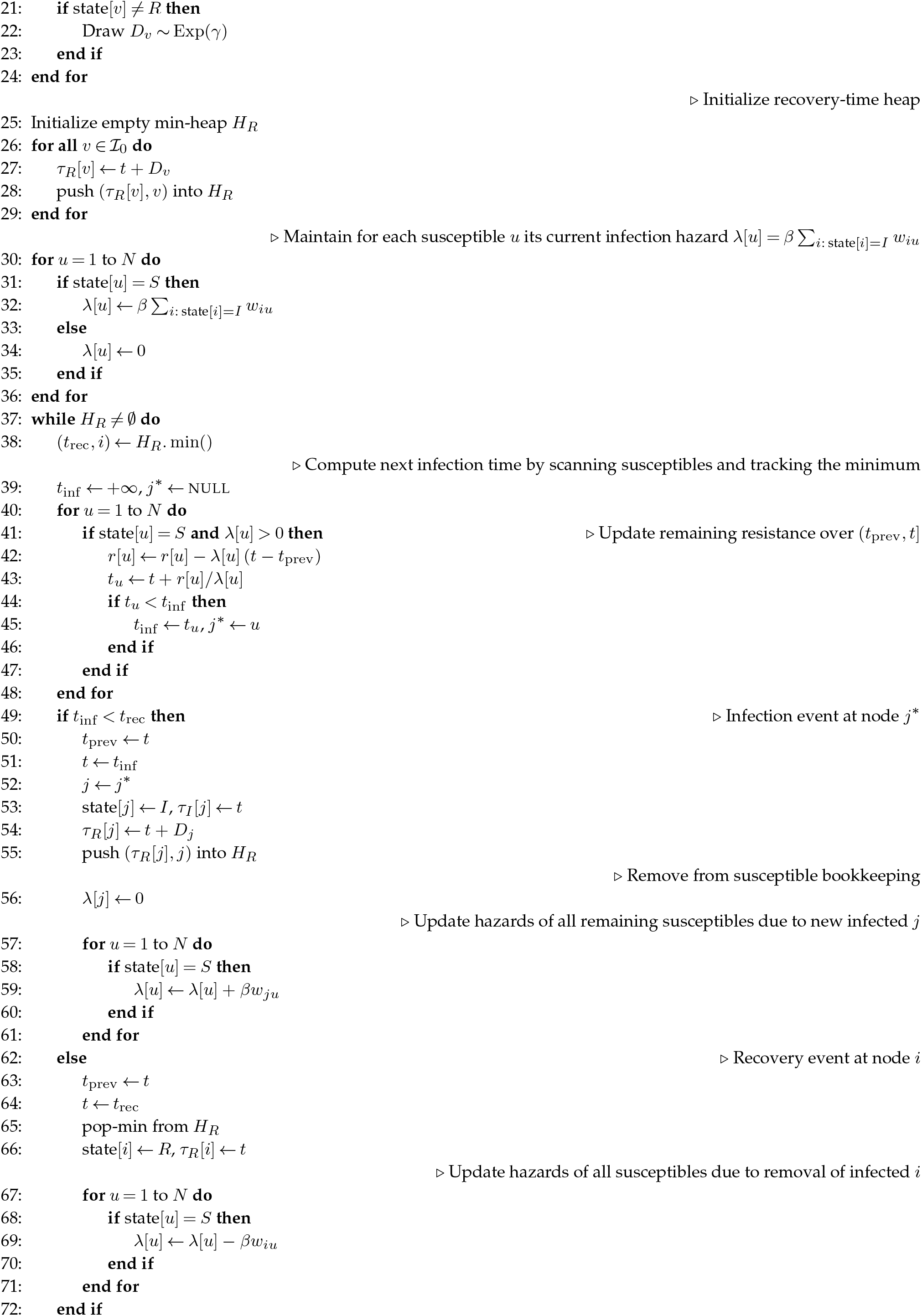
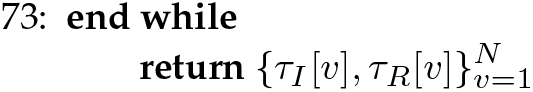

## Notes

### Competing Interest Statement

The authors have declared no competing interest.

